# Epidemiological impact of prioritizing SARS-CoV-2 vaccination by antibody status: Mathematical modeling analyses

**DOI:** 10.1101/2021.01.10.21249382

**Authors:** Houssein H. Ayoub, Hiam Chemaitelly, Monia Makhoul, Zaina Al Kanaani, Einas Al Kuwari, Adeel A. Butt, Peter Coyle, Andrew Jeremijenko, Anvar Hassan Kaleeckal, Ali Nizar Latif, Riyazuddin Mohammad Shaik, Hanan F. Abdul Rahim, Gheyath K. Nasrallah, Hadi M. Yassine, Mohamed G. Al Kuwari, Hamad Eid Al Romaihi, Mohamed H. Al-Thani, Roberto Bertollini, Abdullatif Al Khal, Laith J. Abu Raddad

**Author notes:** **Reprints or correspondence:** Dr. Houssein H. Ayoub, Department of Mathematics, Statistics, and Physics, Qatar University, P.O. Box 2713, Doha, Qatar., Professor Laith J. Abu-Raddad, Infectious Disease Epidemiology Group, World Health Organization Collaborating Centre for Disease Epidemiology Analytics on HIV/AIDS, Sexually Transmitted Infections, and Viral Hepatitis, Weill Cornell Medicine - Qatar, Qatar Foundation - Education City, P.O.Box 24144, Doha, Qatar. **Funding:** The Biomedical Research Program and the Biostatistics, Epidemiology and Biomathematics Research Core at Weill Cornell Medicine-Qatar, Ministry of Public Health, and Hamad Medical Corporation. **Disclose funding received for this work:** others.

## Abstract

**Background:** Vaccines against SARS-CoV-2 have been developed, but their availability falls far short of global needs. This study aimed to investigate the impact of prioritizing available doses on the basis of recipient antibody status, that is by exposure status, using Qatar as an example.

**Methods:** Vaccination impact was assessed under different scale-up scenarios using a deterministic meta-population mathematical model describing SARS-CoV-2 transmission and disease progression in the presence of vaccination.

**Results:** For a vaccine that protects against infection with an efficacy of 95%, half as many vaccinations were needed to avert one infection, disease outcome, or death by prioritizing antibody-negative individuals for vaccination. Prioritization by antibody status reduced incidence at a faster rate and led to faster elimination of infection and return to normalcy. Further prioritization by age group amplified the gains of prioritization by antibody status. Gains from prioritization by antibody status were largest in settings where the proportion of the population already infected at the commencement of vaccination was 30-60%, which is perhaps where most countries will be by the time vaccination programs are up and running. For a vaccine that only protects against disease and not infection, vaccine impact was reduced by half, whether this impact was measured in terms of averted infections or disease outcomes, but the relative gains from using antibody status to prioritize vaccination recipients were similar.

**Conclusions:** Major health, societal, and economic gains can be achieved more quickly by prioritizing those who are antibody-negative while doses of the vaccine remain in short supply.

## INTRODUCTION

The severe acute respiratory syndrome coronavirus 2 (SARS-CoV-2) pandemic has been one of the most challenging global health emergencies in recent history [1, 2]. It is widely believed that vaccination offers the most effective solution to this emergency [3]. More than a hundred vaccines are currently under development [4], with three of them reporting efficacies as high as 95% [5-7], but access to them remains a formidable challenge. Speed of production, logistics, and costs act as barriers for many countries to benefit from vaccine development. With supply limitations and high demand, it is foreseeable that a large proportion of the world’s population may not have access to these vaccines before 2022.

Prioritizing vaccination for specific subpopulations that will benefit most from it is one potential approach to optimize vaccine impact while vaccine supply is being expanded. Evidence suggests that reinfection with SARS-CoV-2 is a rare phenomenon and that most infected persons develop protective immunity against reinfection that lasts for at least a few months post-primary infection [8-10]. Therefore, vaccination is conceivably more beneficial for those who are antibody-negative than those whose immune systems have already confronted this infection and cleared it.

Against this background, the objective of this study was to investigate the impact of vaccination with or without prioritization by antibody status (that is exposure status), using Qatar as an example. With the exact vaccine mechanism of action still unclear, its impact was assessed assuming two possible mechanisms of action, acting against both infection and disease, or acting only against disease. The study was possible thanks to a synergistic application of two innovations in public health systems: use of mathematical modeling to inform public health response, and use of digital healthcare systems to link diverse health information systems, create and analyze databases, and use of outputs for development of mathematical models to forecast the epidemic trajectory, healthcare needs, and impact of interventions such as vaccination.

## METHODS

### Mathematical model

A deterministic meta-population mathematical model was constructed to assess the impact of SARS-CoV-2 vaccination in Qatar by extending and adapting our previously validated and published models [3, 11-13]. The model description is summarized below, and further details can be found in the previous publications, particularly references [3, 11].

The model consisted of a set of coupled, nonlinear differential equations and was structured by age (0-9, 10-19, …, ≥80 years) and grouped by the major nationalities of the population of Qatar. Unvaccinated and vaccinated populations were further stratified based on infection status (uninfected, infected), infection stage (mild, severe, critical), and disease stage (severe, critical) (Figure S1).

Susceptible populations were assumed at risk of acquiring the infection at a hazard rate that varies based on the infectious contact rate per day, nationality, age-specific exposure/susceptibility to the infection, and subpopulation mixing and age group mixing matrices, parametrizing mixing between individuals in different nationality and age groups. Infected individuals develop mild (or asymptomatic), severe, or critical infections, following a latency period. The proportion of infected persons developing mild, severe, or critical infections was age-dependent, based on relative risks that were based on the SARS-CoV-2 epidemic in France [14]. Severe and critical infections progress to severe and critical disease, respectively, prior to recovery. These are hospitalized in acute-care and ICU-care beds, respectively, based on existing standards of care. Critical disease cases have an additional risk of COVID-19 mortality. The model was coded, fitted, and analyzed using MATLAB R2019a [15].

### Model parametrization and fitting

Model parameterization was based on current data for SARS-CoV-2 natural history and epidemiology. The model was calibrated through fitting to the standardized and centralized databases of SARS-CoV-2 testing, infections, hospitalizations, and mortality in Qatar [16, 17], as well as to findings of recently-completed epidemiologic studies [16, 18-20]. Fitting to input data was performed using a nonlinear least square fitting technique, based on the Nelder-Mead simplex algorithm.

### Characteristics of the novel vaccine and its scale-up

Since the primary end point of vaccine randomized clinical trials was efficacy against laboratory-confirmed COVID-19 cases [6, 7, 21], and not *any* infection documented or undocumented, it is unknown whether the vaccine acted by prophylactically reducing susceptibility to the infection (that is *VE*_*S*_ efficacy, defined as the proportional reduction in susceptibility to infection among those vaccinated, compared to those unvaccinated [3]), or whether it simply acted by reducing serious symptomatic COVID-19 cases with no effect on infection (that is *VE*_*P*_ efficacy against disease progression, defined as a proportional reduction in the fraction of individuals with severe or critical infection among those vaccinated, but who still acquired the infection, compared to those unvaccinated [3]). These two mechanisms of action bracket the two extremes for the vaccine’s biological effect and impact, with the reduction of both infection and disease being the most optimistic and the reduction of only severe disease forms being the most conservative. Notwithstanding this uncertainty, the impact of the vaccine was assessed assuming each of these mechanisms of action, *VE*_*S*_ = 95% and *VE*_*P*_ = 95%, and assuming that the vaccine will offer one year of protection.

### Vaccine program scenarios

Several vaccination scenarios were considered and these were informed by the availability of the vaccine in Qatar and the tentative schedule of its incoming shipments over the coming months. The first shipment of the Pfizer-BioNTech COVID-19 vaccine arrived on December 21, 2020, and vaccination has just been launched.

The considered vaccination scenarios included administering the vaccine only to those who are antibody-negative, or irrespective of antibody status, administering a specific number of vaccinations or vaccinating to reach a specific coverage in a specific target population, and prioritizing specific age brackets as opposed to others. While the impact of vaccination in Qatar was the focus of this study, the generic impact of vaccination was also assessed at *different* assumed levels of infection exposure in the population at time of onset of vaccination, to reflect generically the diversity of the epidemic situation in different countries.

It was assumed that the vaccine will be introduced in January 1, 2021 and will be scaled up within six months. Since the purpose of vaccination is to alleviate the need for restrictions that affect social and economic activities, it was assumed that social and physical distancing restrictions will be eased gradually during these six months, so that full “normalcy” will be attained. Normalcy was defined as a contact rate in the population that is similar to that prior to the pandemic, leading to a basic reproduction number *R*_0_ = 4 at the end of the six-month duration for easing of restrictions. The value of *R*_0_ = 4 is justified by existing estimates of *R*_0_ for an epidemic in absence of interventions [22, 23].

### Measures of vaccine impact

Direct and indirect public health benefits of vaccination were assessed. The direct impact results from direct effects of the vaccine (*VE*_*S*_ or *VE*_*P*_). The indirect impact results from the reduction in onward transmission of the infection, applicable only in the case of *VE*_*S*_.

The total impact of the vaccine, the sum of its direct and indirect impacts, was estimated by comparing incidence at a given time in presence of vaccination, with that in the no-vaccination counter-factual scenario. Impact was also estimated by quantifying *effectiveness*, the number of vaccinations needed to avert one infection or one adverse disease outcome during a specific period. This metric is essentially cost-effectiveness, but with no costs included. Impact of the vaccine was further assessed by estimating the number of days needed to eliminate the infection after initiating vaccination, with infection elimination being defined as an incidence rate ≤1 infection per 100,000 person-days.

### Uncertainty analyses

Ranges of outcome uncertainty predicted by the model were calculated using five-hundred simulation runs that applied Latin Hypercube sampling [24, 25] from a multidimensional distribution of model parameters. At each run, input parameter values were selected from ranges specified by assuming ±30% uncertainty around parameter point estimates. The resulting distribution for each outcome predicted by the model was then used to derive the means and associated 95% uncertainty intervals (UIs) for vaccine effectiveness at each time point. Further details about this type of uncertainty analysis can be found in [11].

## RESULTS

For 500,000 vaccinations administered in the first six months of the year (*VE*_*S*_ = 95%), vaccination of only antibody-negative persons would yield, by June 30, 2021, a reduction of 98% in the daily number of new infections, 83,200 averted infections, 5.9 vaccinations to avert one infection, and 155 days to eliminate the infection (Figure 1). Meanwhile, vaccination irrespective of antibody status would yield, by June 30, 2021, a reduction of 73% in the daily number of new infections, 40,600 averted infections, 12.0 vaccinations to avert one infection, and 228 days to eliminate the infection.

**Figure 1:**
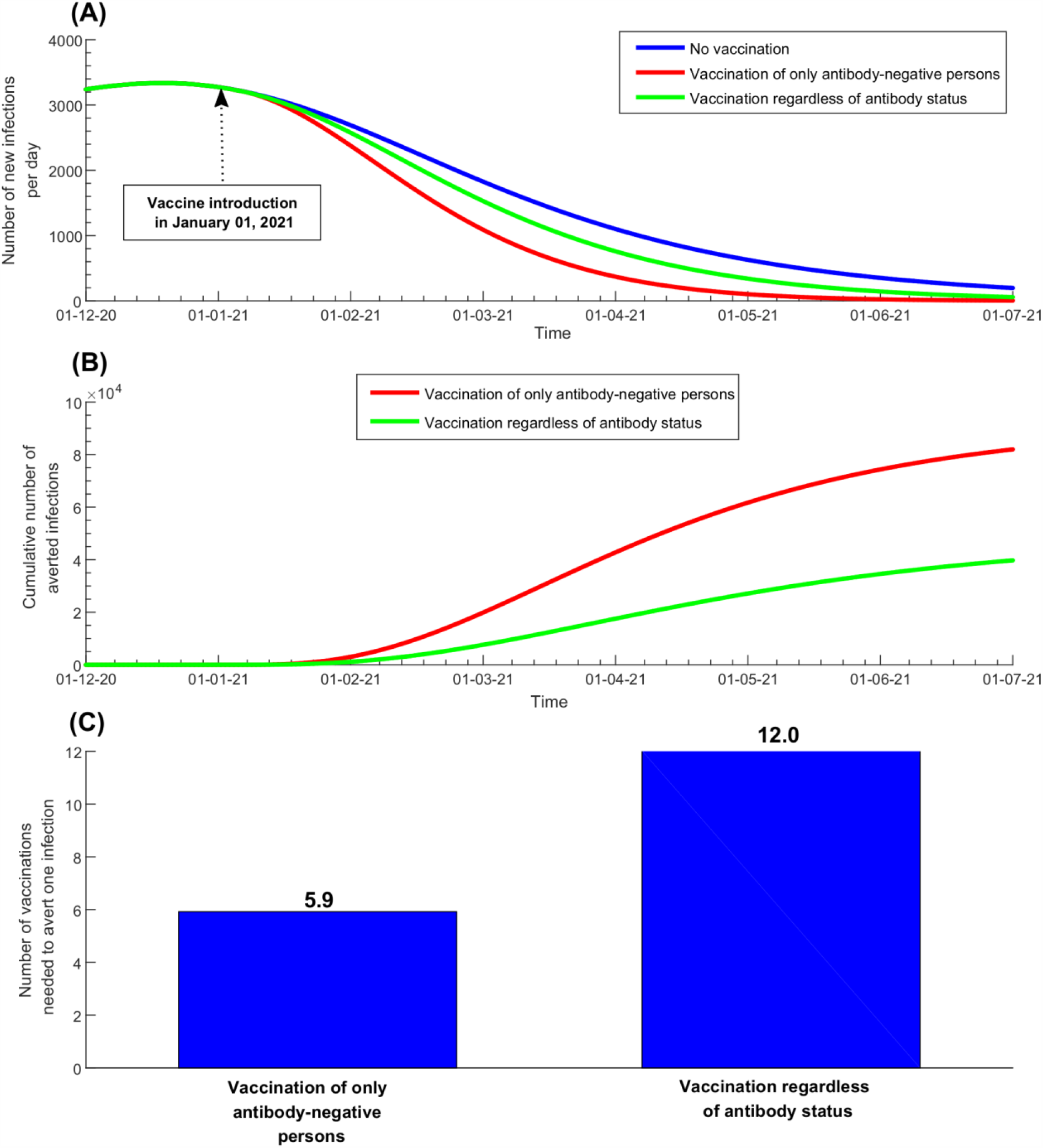
Impact of 500,000 SARS-CoV-2 vaccinations with or without prioritization by antibody status. Impact was assessed based upon A) the number of new infections, B) the cumulative number of averted infections, and C) the number of vaccinations needed to prevent one infection. Vaccination is introduced on January 1^st^, 2021 and is scaled up until June 30, 2021, with concurrent gradual easing of social and physical distancing restrictions to reach an *R*_0_ of 4 by June 30, 2021. The vaccine is assumed to have an efficacy of 95% against infection: *VE*_*S*_ = 95%. Duration of vaccine-induced protection is one year.

For *VE*_*S*_ = 95%, Figure 2 shows the impact of achieving vaccine coverage of 80% only among those who are antibody-negative, or of reaching 80% coverage in the whole population, by June 30, 2021. As expected, the impact of the vaccine on infection is the same in both scenarios, as the number of people who benefited from the vaccine (only those antibody-negative) is the same in both scenarios. Seventy-seven days are needed to reach elimination, but elimination is reached with far fewer vaccinations if only those who are antibody-negative are prioritized. This is reflected in effectiveness, as only 8.6 vaccinations would be needed to avert one infection by prioritizing antibody-negative persons, but 20.6 vaccinations would be needed by vaccinating irrespective of antibody status. Similar results are found for gains attained by prioritizing according to antibody status in the case of a vaccine that only reduces disease with *VE*_*P*_ = 95% (Figure S2).

**Figure 2:**
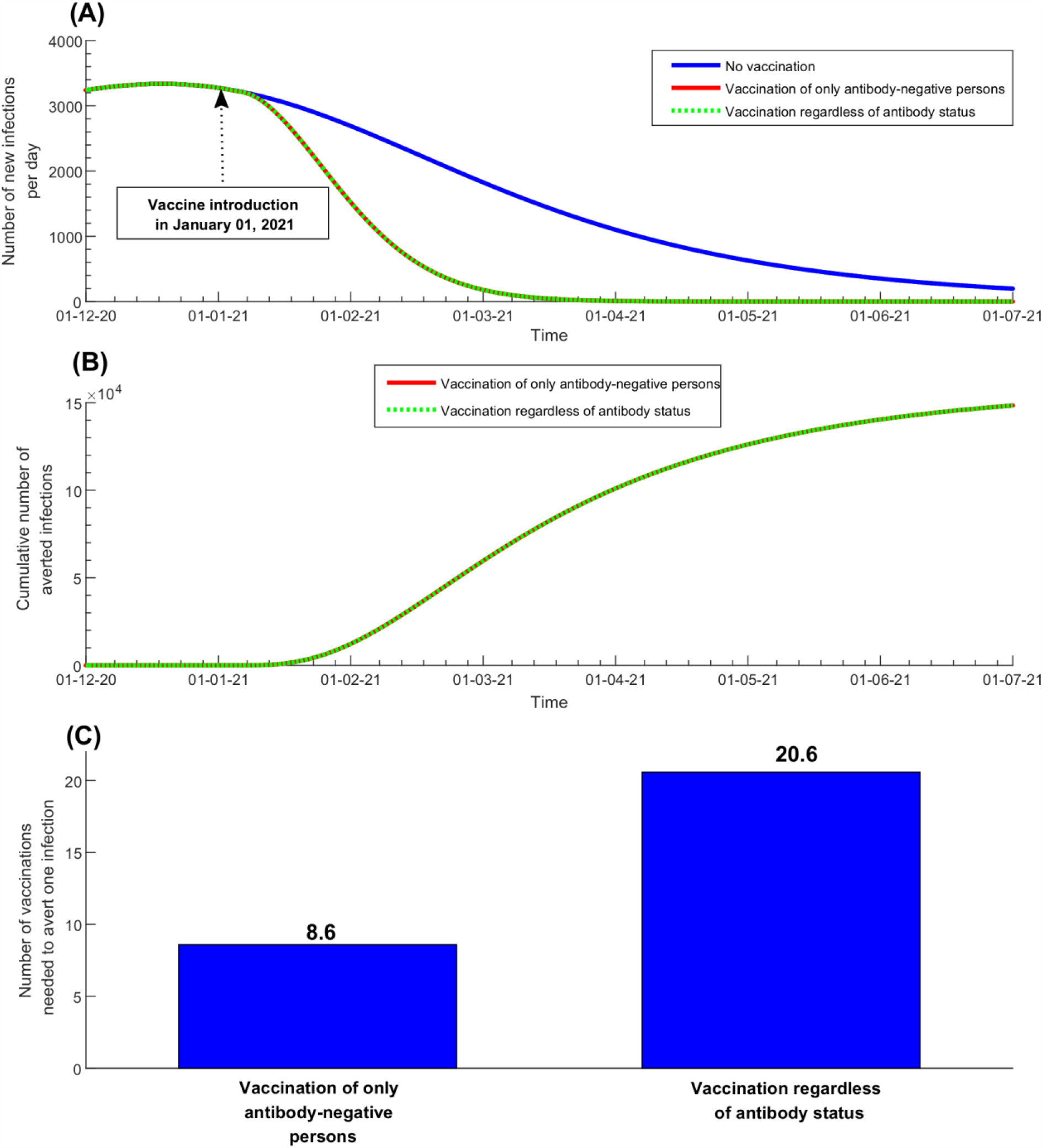
Impact of SARS-CoV-2 vaccination to reach 80% coverage among only the antibody-negative, or to reach 80% coverage of the whole population. Impact was assessed based upon A) the number of new infections, B) the cumulative number of averted infections, and C) the number of vaccinations needed to prevent one infection. Vaccination is introduced on January 1^st^, 2021 and is scaled up until June 30, 2021, with concurrent gradual easing of social and physical distancing restrictions to reach an *R*_0_ of 4 by June 30, 2021. The vaccine is assumed to have an efficacy of 95% against infection: *VE*_*S*_= 95%. Duration of vaccine-induced protection is one year.

Figure 3 shows the impact of SARS-CoV-2 vaccination to reach 80% coverage among those antibody-negative for a vaccine that reduces both infection and disease (*VE*_*S*_ = 95%) compared to a vaccine that reduces only disease (*VE*_*P*_ = 95%). Figure 4 shows the corresponding effectiveness in terms of the number of vaccinations needed to avert one severe disease case, one critical disease case, or one COVID-19 death. A vaccine with *VE*_*S*_ = 95% has a two-fold higher impact than a vaccine with *VE*_*P*_ = 95%, whether this impact is measured in terms of averted infections or disease outcomes (Figure 3), or effectiveness in terms of the number of vaccinations needed to avert one disease outcome (Figure 4).

**Figure 3:**
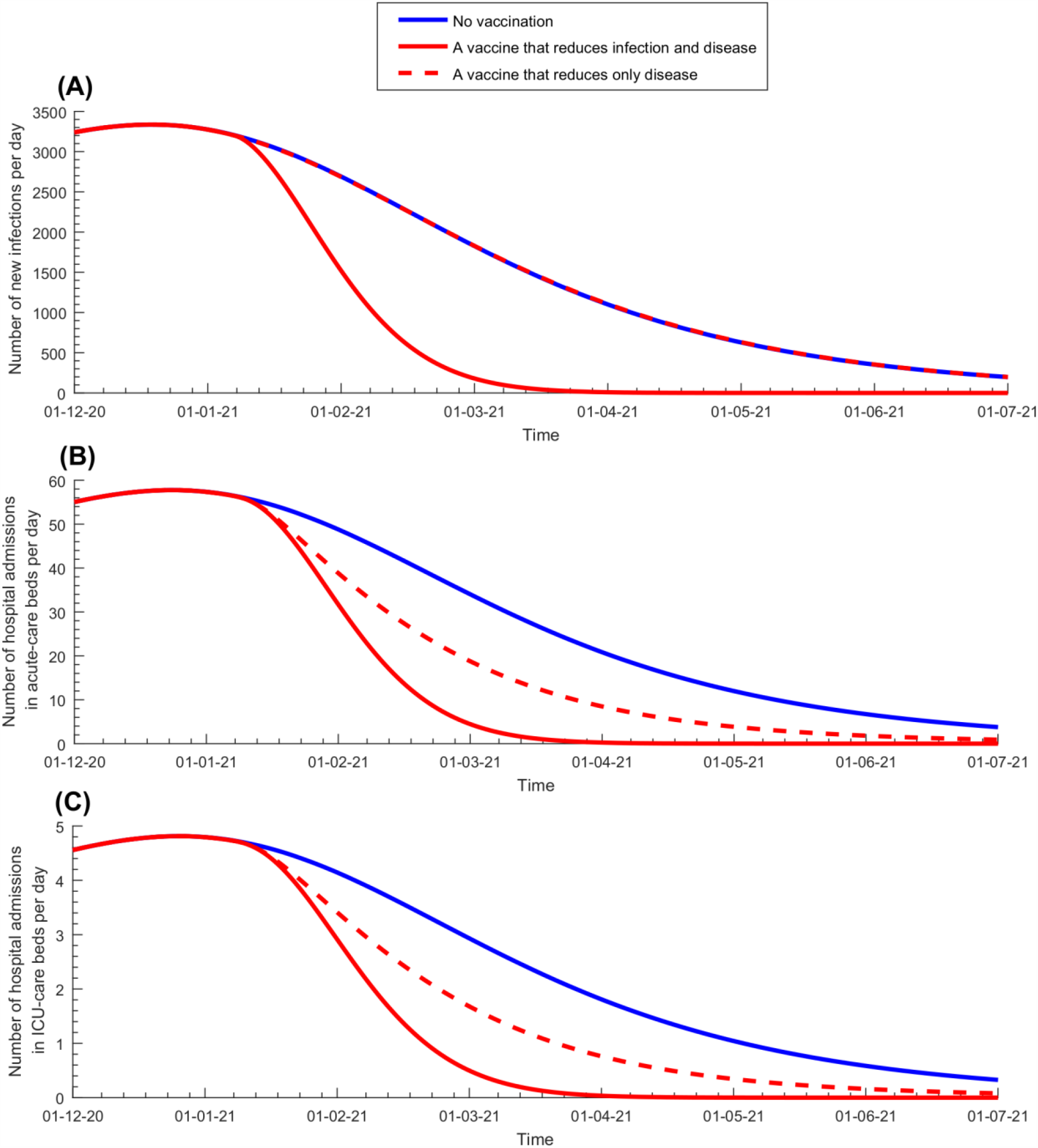
Impact of SARS-CoV-2 vaccination to reach 80% coverage for a vaccine that reduces infection and disease (*VE*_*S*_ = 95%) compared to a vaccine that reduces only disease (*VE*_*P*_ = 95%). Impact was assessed based upon A) the number of new infections per day, B) the number of new hospital admissions in acute-care beds per day, and C) the number of new hospital admissions in ICU beds per day. Only those who are antibody-negative are being vaccinated. Vaccination is introduced on January 1^st^, 2021 and is scaled up until June 30, 2021, with concurrent gradual easing of social and physical distancing restrictions to reach an *R*_0_ of 4 by June 30, 2021. Duration of vaccine-induced protection is one year.

**Figure 4:**
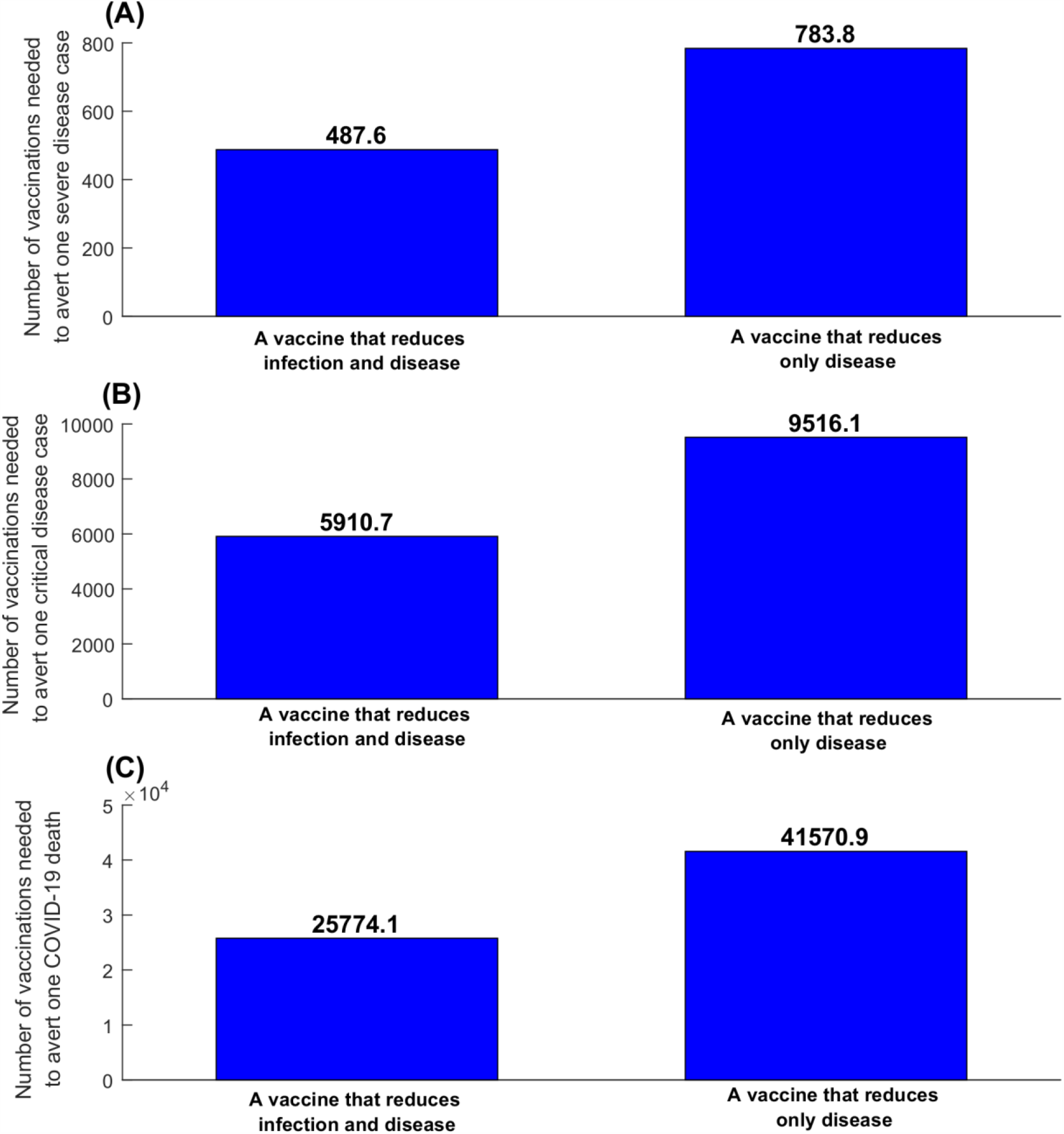
Effectiveness of SARS-CoV-2 vaccination for a vaccine that reduces infection and disease (*VE*_*S*_ = 95%) compared to a vaccine that reduces only disease (*VE*_*P*_ = 95%). The number of vaccinations needed to prevent A) one severe disease case, B) one critical disease case, and C) one COVID-19 death. Only those antibody-negative are being vaccinated with a coverage of 80%. Vaccination is introduced on January 1^st^, 2021 and is scaled up until June 30, 2021, with concurrent gradual easing of social and physical distancing restrictions to reach an *R*_0_ of 4 by June 30, 2021. Duration of vaccine-induced protection is one year.

Figure S3 shows, for *VE*_*S*_ = 95%, the effectiveness of age-group prioritization in administering the vaccine only to those who are antibody-negative. Fewer vaccinations would be needed to avert one infection or one disease outcome by prioritizing the vaccine for those 20-49 years of age and older, as expected given the lower susceptibility to infection for children as opposed to adults. Figure S4 shows the same results, but by administering the vaccine irrespective of antibody status. While vaccinating those 20-49 years of age and older irrespective of antibody status is also more effective, the differential gains are reduced and the effectiveness has a more complex pattern. This complexity arises from the fact that seroprevalence varies considerably by age in Qatar with the lowest levels among children, followed by those >50 years of age, and is highest among those 20-49 years of age [16, 19, 20, 26].

The above results show the impact of vaccination in Qatar, a country where 56.2% of the population is estimated, through serological surveys and mathematical modeling [11, 16, 19, 20, 26], to have been infected by January 1, 2021, at the onset of vaccination. Meanwhile, Figure 5 shows the impact of vaccination at *different* assumed levels of infection exposure in the population at the onset of vaccination. The figure specifically compares the number of days needed to eliminate the infection in a scenario in which vaccination is administered only to people antibody-negative at a coverage of 80%, to a scenario in which an *equal number* of vaccinations was administered, but irrespective of antibody status. In the scenario in which only those antibody-negative are being vaccinated, the higher the infection exposure is at onset of vaccination, the less time is needed to reach elimination, as expected, as the vaccine is provided *only* to those who will directly benefit from it.

**Figure 5:**
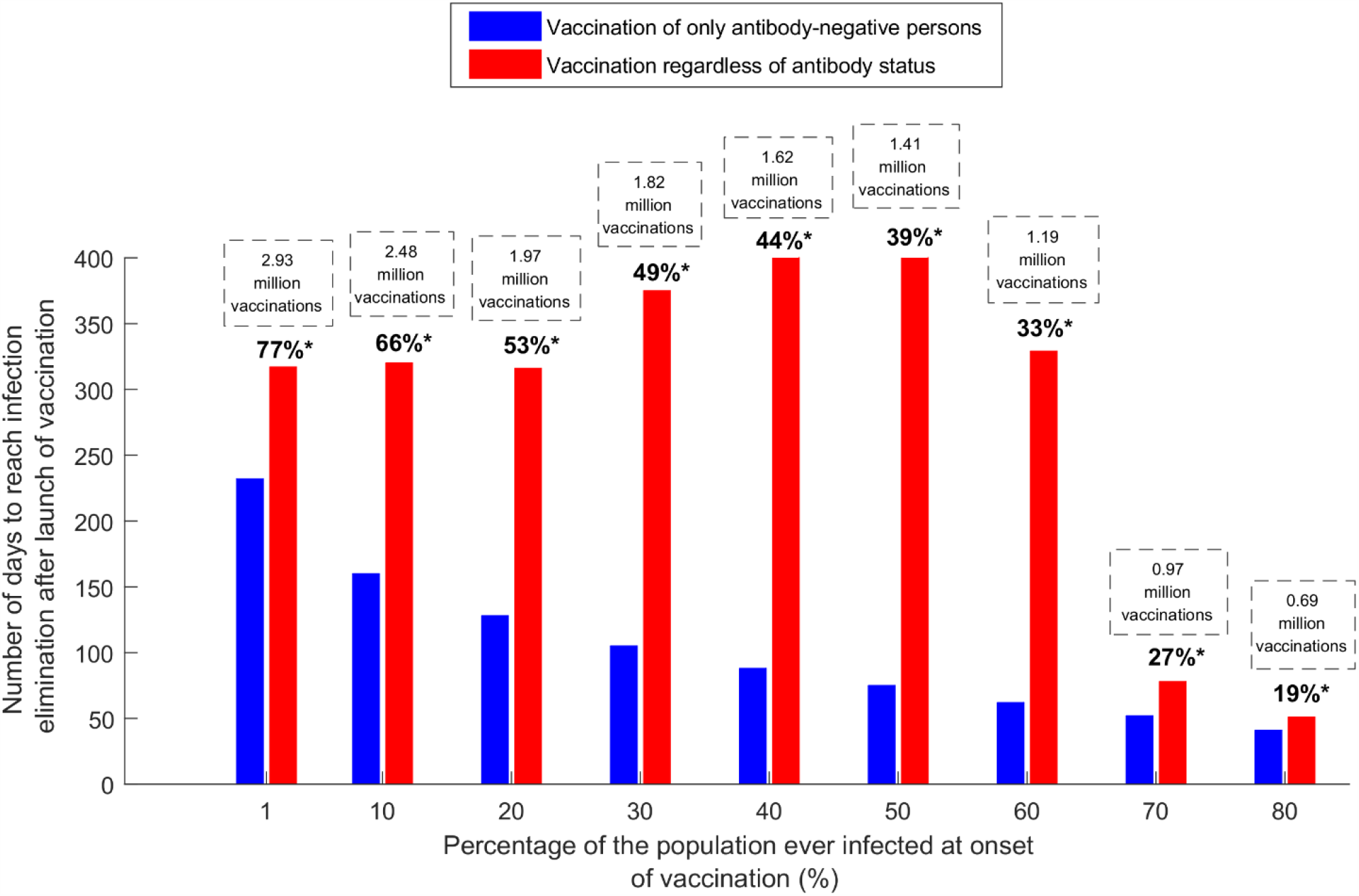
The number of days needed to eliminate the infection after launching vaccination at different assumed levels of infection exposure (attack rate) in the population at time of vaccination onset. The number of days needed to eliminate the infection in a scenario in which vaccination is administered only to those who are antibody-negative at 80% coverage, is compared to a scenario in which an *equal* number of vaccinations was administered, but irrespective of antibody status. Vaccination is introduced on January 1^st^, 2021 and is scaled up until June 30, 2021, with concurrent gradual easing of social and physical distancing restrictions to reach an *R*_0_ of 4 by June 30, 2021. The vaccine is assumed to have an efficacy of 95% against infection: *VE*_*S*_ = 95%. Duration of vaccine-induced protection is one year.

However, the situation is more nuanced for the scenario in which individuals are vaccinated irrespective of antibody status. If infection exposure is very low at the onset of vaccination, less time would be needed to reach elimination, as the vast majority of those vaccinated are antibody-negative and will directly benefit from the vaccine. If infection exposure is very high at onset of vaccination (>60%), less time would also be needed to reach elimination, as the population is already close to the herd immunity threshold (at 80% infection exposure for *R*_0_ of 4), and will attain it quickly, even though most of those vaccinated are already antibody-positive and will not directly benefit from vaccination. The longest time to elimination is seen when infection exposure at onset of vaccination is in the intermediate range, between 30-60%, as the population is not close to the herd immunity threshold, but at the same time, many of those vaccinated have already been exposed to the infection and will not directly benefit from the vaccine.

Figure S5 shows the results of the uncertainty analysis for vaccine effectiveness. The results demonstrate relatively narrow uncertainty intervals, thereby affirming the results.

## DISCUSSION

The first finding of this study is that there are major gains by prioritizing available vaccines to persons who are antibody-negative, regardless of whether the vaccine reduces infection and disease, or just disease. With vaccine availability falling far short of global needs, such prioritization will reduce the incidence rate of the infection more quickly, thereby eliminating the infection and returning to normalcy sooner. Vaccination would thus avert more disease cases and deaths and would be more cost-effective, with fewer vaccinations needed to avert one infection or disease outcome. As much as our results point toward substantial health, societal, and economic gains for vaccine prioritization by exposure status, actual implementation of such an approach is still contingent on the feasibility and cost of wide-scale antibody testing, as a component of vaccination programs in various countries, as well as equity in prioritizing the vaccine for some as opposed to others.

The second finding of this study is that the gains of prioritizing vaccination by antibody status are largest in settings where the proportion of the population previously infected (at time of launch of vaccination) is between 30-60%, which is perhaps where most countries will be by the time vaccination programs are up and running, affirming major dividends to be reaped from this approach. For countries that are still at limited infection exposure, prioritization by antibody status will not yield such significant gains, as very few vaccinations are given to those previously infected, irrespective of whether prioritization is implemented.

A third finding of this study is that the impact of the vaccine depends on whether the vaccine reduces infection and disease, or reduces only disease. The impact of the former was two-fold higher than the impact of the latter, regardless of whether this impact is measured in terms of averted disease cases, or in terms of the number of vaccinations needed to avert one disease outcome. This finding is explained by the fact that for a vaccine that reduces susceptibility to infection (a “*VE*_*S*_” vaccine), half of the beneficial impact is *indirect*, by reducing the onward transmission of the infection in the population, in addition to the *direct* impact of preventing infection among those vaccinated.

This study has some limitations. Model estimates are contingent on the validity and generalizability of input data and assumptions. Our results are based on current understanding of SARS-CoV-2 natural history and disease progression, but our understanding of this infection is still evolving. A key assumption is that those infected acquire protective immunity against reinfection that lasts for at least a year. While this assumption is supported by current evidence [8-10], studies with longer-term follow-up are still needed to assess the duration of natural immunity. Vaccine-induced immunity is assumed to last for one year, but the duration of this immunity is also unknown. Therefore, model predictions may not be valid if either duration of natural immunity or vaccine-induced immunity lasts less than a year, whether because of waning immunity or appearance of mutant virus strains that escape immunity. The model assumes that vaccinated persons are protected immediately once vaccinated, but vaccine protection builds up gradually over the course of a month following inoculation and peaks after the second does [6]. This may slightly reduce the gains projected here.

In conclusion, major health, societal, and economic gains can be attained by prioritizing vaccination for those who are antibody-negative, as long as doses of the vaccine remain in short supply.

## Data Availability

All data are available within the manuscript and its supplementary materials.

## Acknowledgements

We thank Her Excellency Dr. Hanan Al Kuwari, Minister of Public Health, for her vision, guidance, leadership, and support. We also thank Dr. Saad Al Kaabi, Chair of the System Wide Incident Command and Control (SWICC) Committee for the COVID-19 national healthcare response, for his leadership, analytical insights, and for his instrumental role in enacting data information systems that made these studies possible. We further extend our appreciation to the SWICC Committee and the Scientific Reference and Research Taskforce (SRRT) members for their informative input, scientific technical advice, and enriching discussions. We also thank Dr. Mariam Abdulmalik, CEO of the Primary Health Care Corporation and the Chairperson of the Tactical Community Command Group on COVID-19, as well as members of this committee, for providing support to the teams that provided field surveillance. We further thank Dr. Nahla Afifi, Director of Qatar Biobank (QBB), Ms. Tasneem Al-Hamad, Ms. Eiman Al-Khayat and the rest of the QBB team for their unwavering support in retrieving and analyzing samples and in compiling and generating databases for COVID-19 infection, as well as Dr. Asmaa Al-Thani, Chairperson of the Qatar Genome Programme Committee and Board Vice Chairperson of QBB, for her leadership of this effort. We also acknowledge the dedicated efforts of the Clinical Coding Team and the COVID-19 Mortality Review Team, both at Hamad Medical Corporation, and the Surveillance Team at the Ministry of Public Health.

## Author contributions

HHA co-designed the study, constructed and parameterized the mathematical model, conducted the mathematical modeling analyses, and co-wrote the first draft of the manuscript. HC conducted the statistical analyses and contributed to the parameterization of the mathematical model. LJA conceived and co-designed the study, led the conduct of the analyses, and co-wrote the first draft of the manuscript. All authors contributed to conceptualization of the analyses, discussion and interpretation of the results, and writing of the manuscript. All authors have read and approved the final manuscript.

## Supporting Information

**Figure S1:**
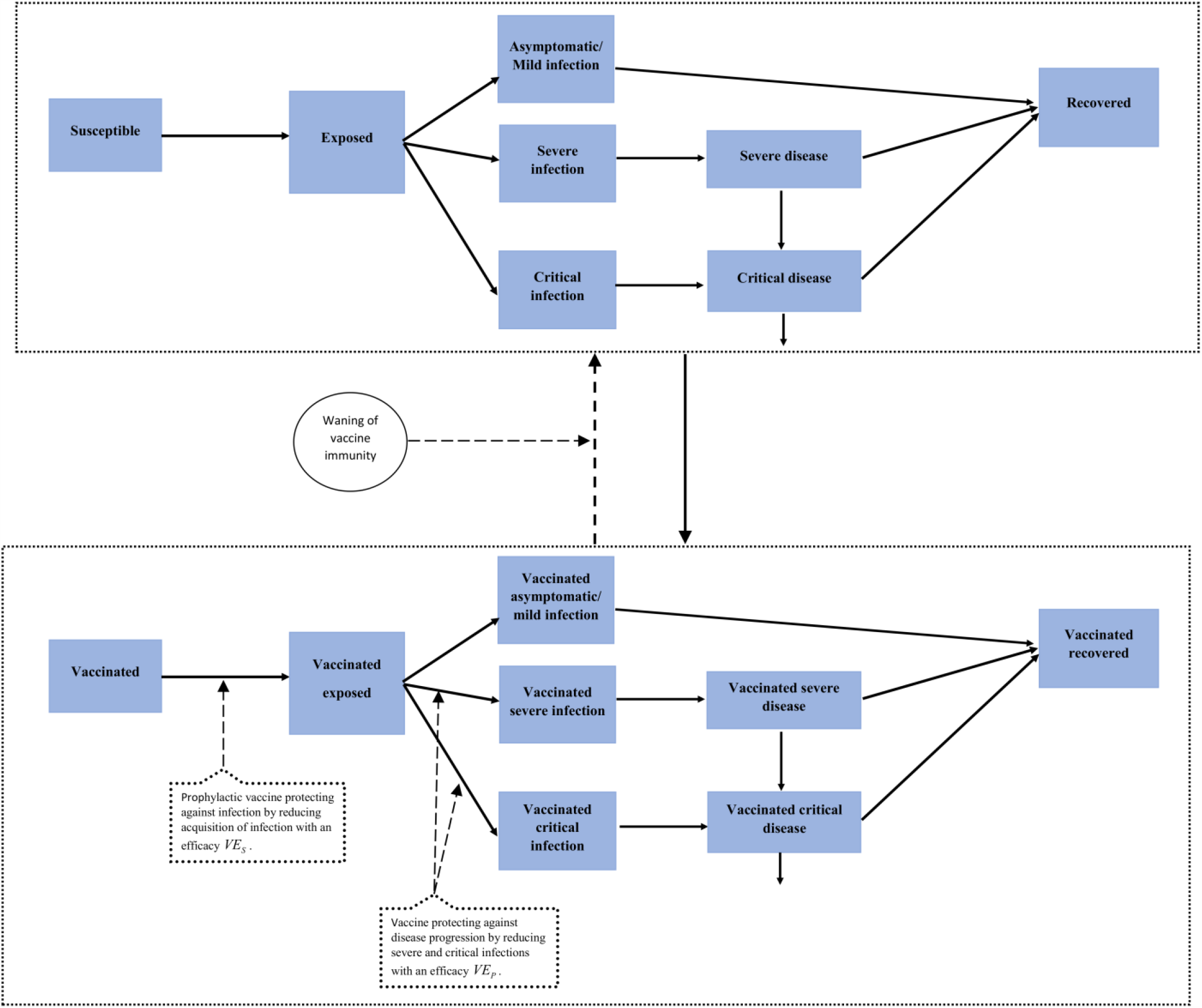
Conceptual diagram illustrating the SARS-CoV-2 vaccine model. *VE*_*S*_ is defined as the proportional reduction in the susceptibility to infection among those vaccinated compared to those unvaccinated.[1] *VE*_*P*_ is defined as the proportional reduction in the proportion of individuals with severe or critical infection among those vaccinated but still acquired the infection compared to those unvaccinated.[1] In this figure, solid lines denote progression or forward movement from one population compartment to the next, while dashed lines denote backward movement from the present population compartment to the previous population compartment. Further details can be found in references.[1-4]

**Figure S2:**
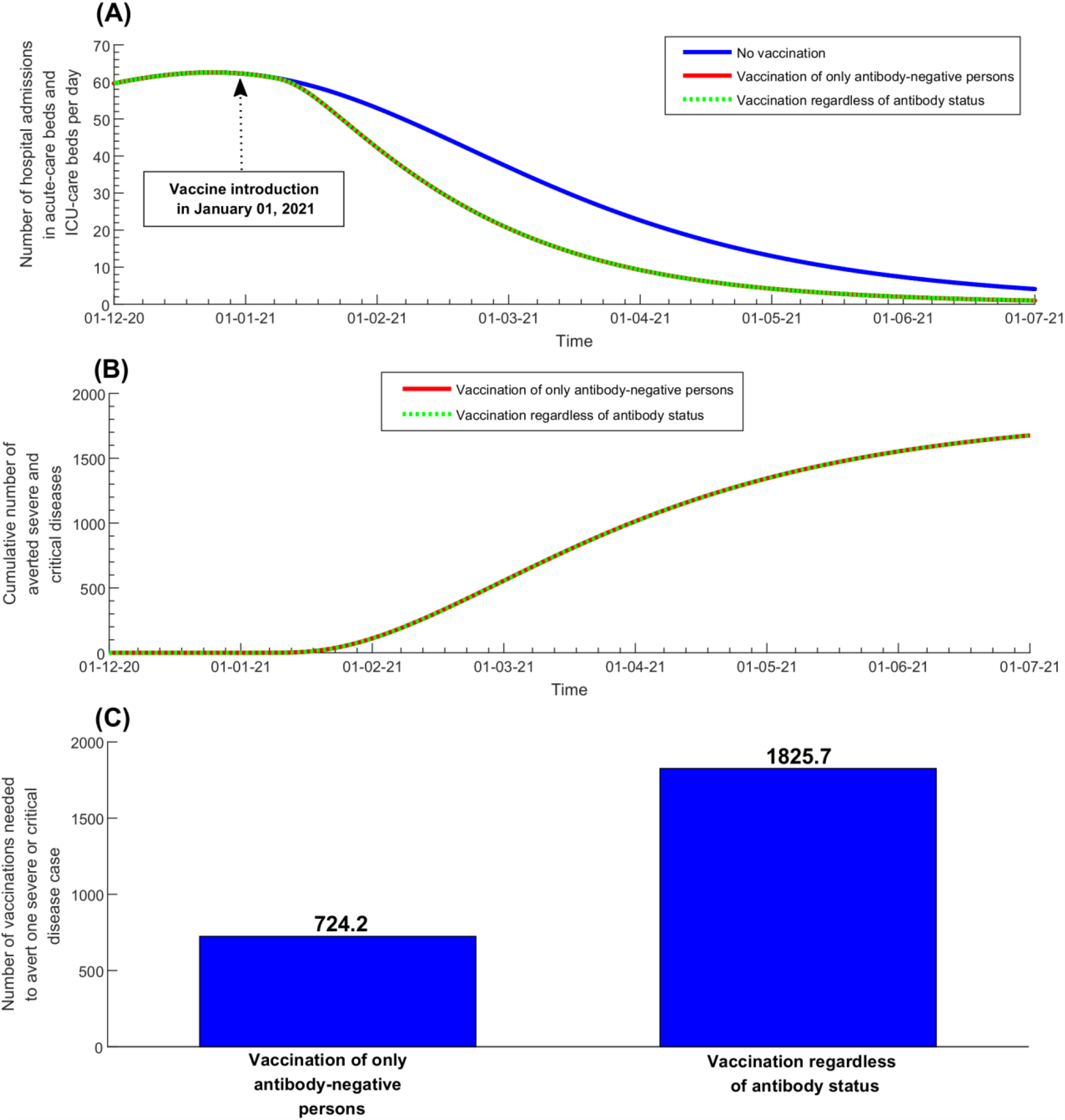
Impact of SARS-CoV-2 vaccination to reach 80% coverage among only the antibody-negative, or to reach 80% coverage of the whole population, for a vaccine that does not protect against infection, but protects against disease. Impact was assessed based upon A) the number of new hospital admissions in acute-care beds and ICU-care beds per day, the cumulative number of averted severe and critical diseases, and C) the number of vaccinations needed to prevent one severe or critical disease case. Vaccination is introduced on January 1^st^, 2021 and is scaled up until June 30, 2021, with concurrent gradual easing of social and physical distancing restrictions to reach an *R*_0_ of 4 by June 30, 2021. The vaccine is assumed to have an efficacy of 95% against only disease: *VE*_*P*_ = 95%. Duration of vaccine-induced protection is one year.

**Figure S3:**
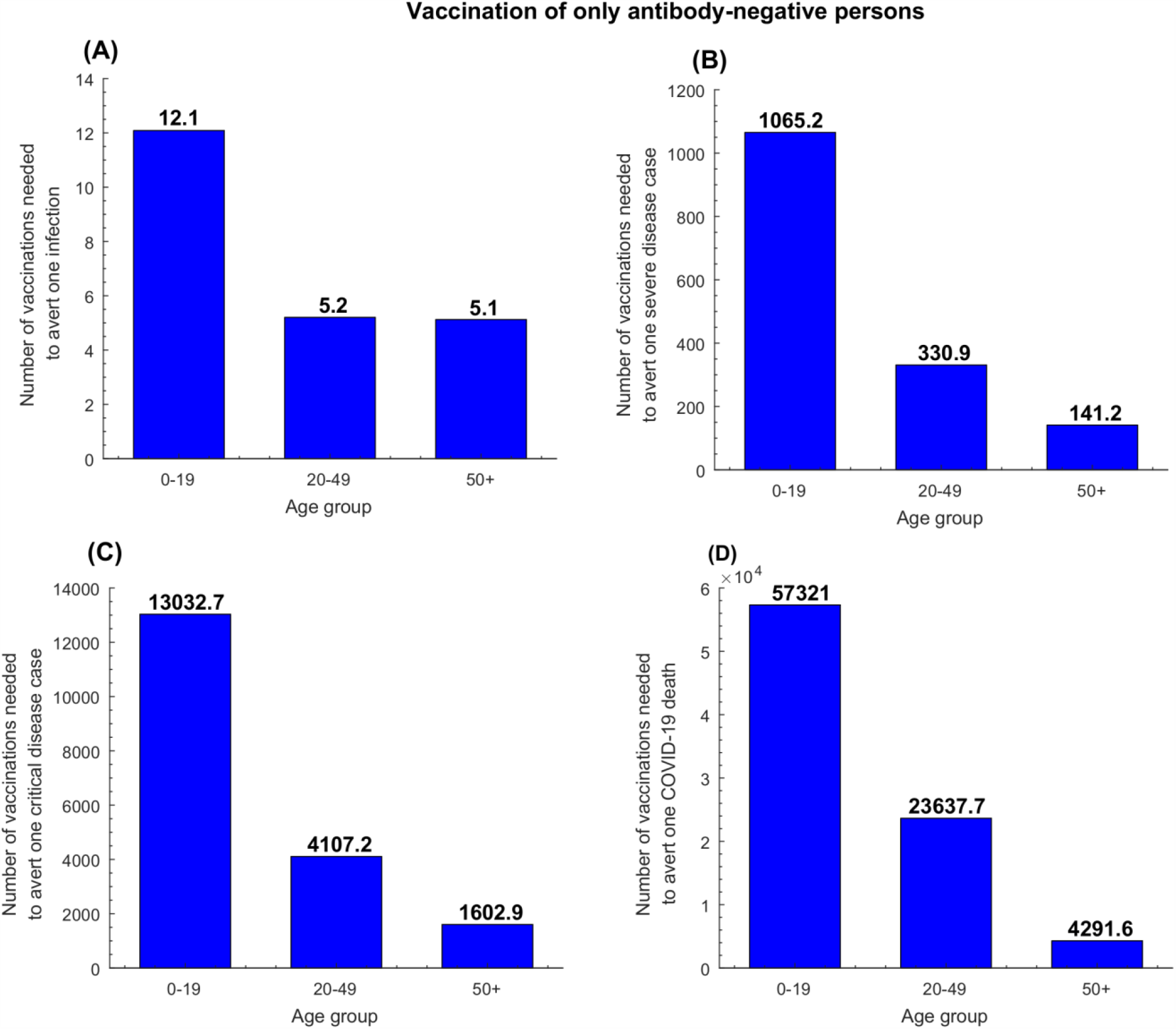
Effectiveness of age-group prioritization in vaccinating only antibody-negative persons. The number of vaccinations needed to prevent A) one infection, B) one severe disease case, C) one critical disease case, and D) one COVID-19 death. Vaccination is introduced on January 1^st^, 2021 and is scaled up until June 30, 2021, with concurrent gradual easing of social and physical distancing restrictions to reach an *R*_0_ of 4 by June 30, 2021. The vaccine is assumed to have an efficacy of 95% against infection: *VE*_*S*_ = 95%. Duration of vaccine-induced protection is one year.

**Figure S4:**
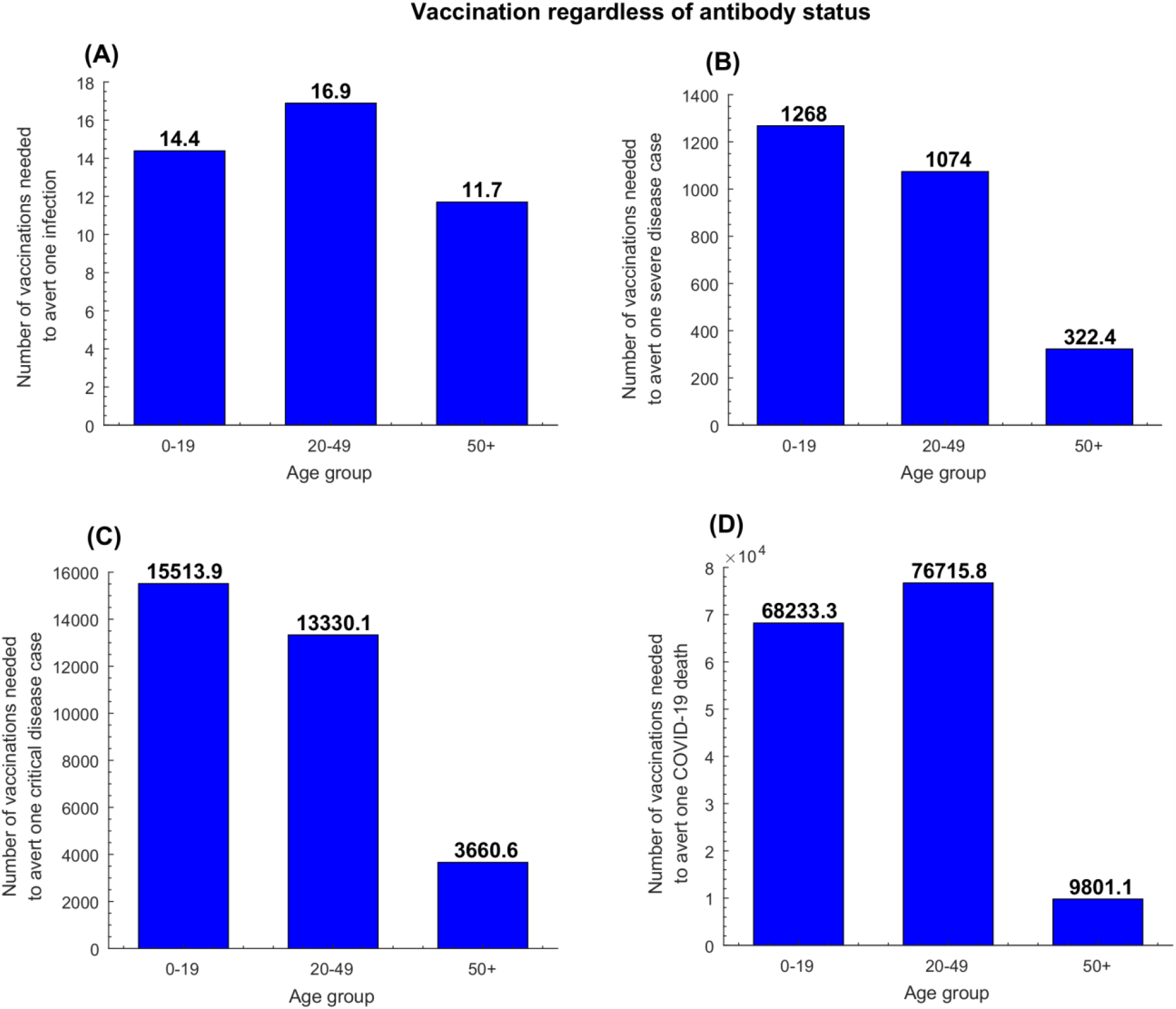
Effectiveness of age-group prioritization in vaccinating regardless of antibody status. The number of vaccinations needed to avert A) one infection, B) one severe disease case, C) one critical disease case, and D) one COVID-19 death. Vaccination is introduced on January 1^st^, 2021 and is scaled up until June 30, 2021, with concurrent gradual easing of social and physical distancing restrictions to reach an *R*_0_ of 4 by June 30, 2021. The vaccine is assumed to have an efficacy of 95% against infection: *VE*_*S*_ = 95%. Duration of vaccine-induced protection is one year.

**Figure S5:**
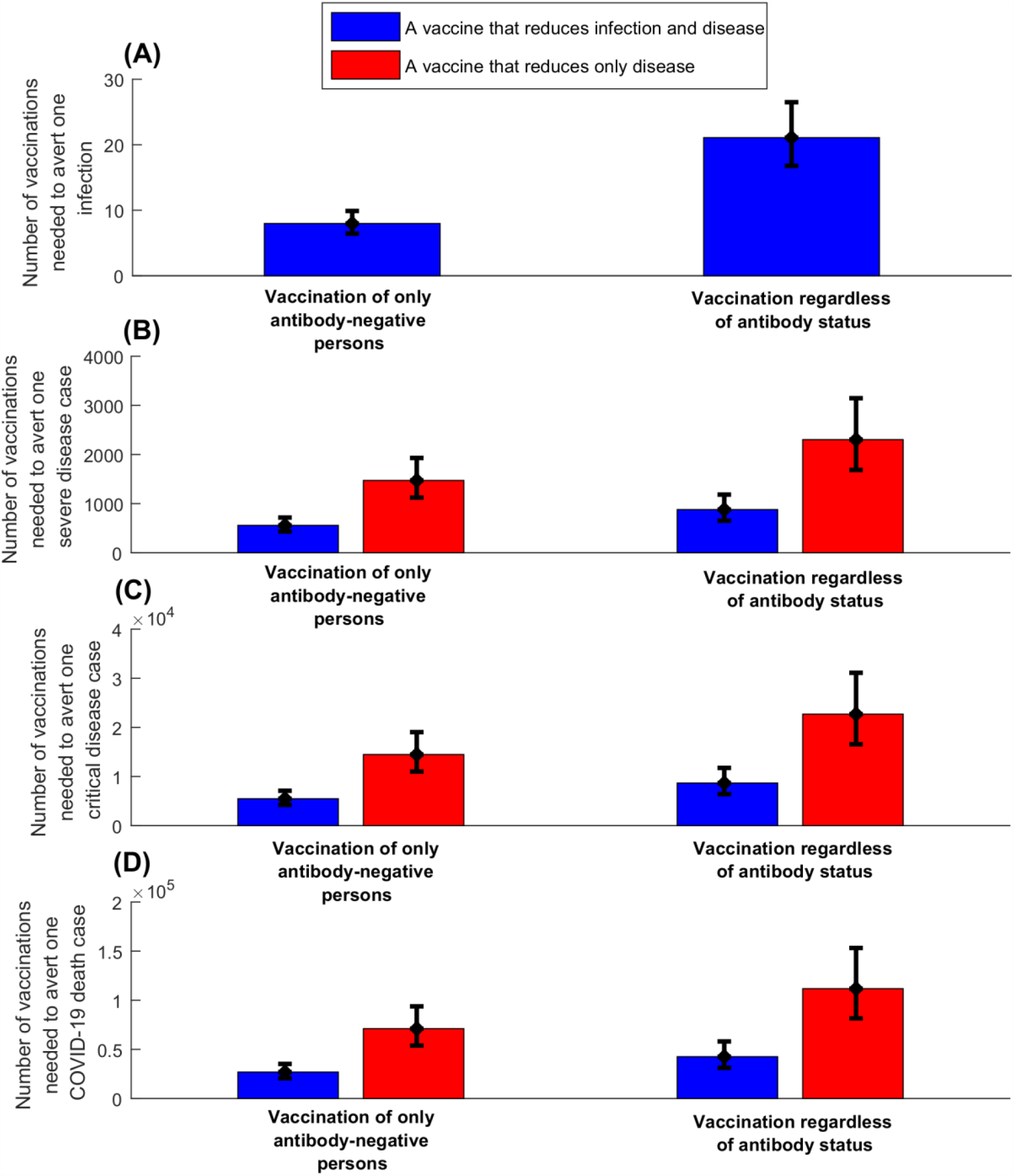
Uncertainty analysis. The mean and 95% uncertainty interval (UI) for the effectiveness of SARS-CoV-2 vaccination with or with no prioritization by antibody status for a vaccine that reduces infection and disease (*VE*_*S*_ = 95%) compared to a vaccine that reduces only disease (*VE*_*P*_ = 95%). The number of vaccinations needed to avert A) one infection A) one severe disease case, B) one critical disease case, and C) one COVID-19 death. Vaccination is introduced on January 1^st^, 2021 and is scaled up until June 30, 2021, with concurrent gradual easing of social and physical distancing restrictions to reach an *R*_0_ of 4 by June 30, 2021. Duration of vaccine-induced protection is one year. of 4 by June 30, 2021.

